# Post-traumatic stress disorder and REM-sleep behavior disorder: exploring genetic associations and causal links

**DOI:** 10.1101/2025.09.05.25335205

**Authors:** Morvarid Ghamgosar Shahkhali, Lang Liu, Mohammad H. Ghamgosar Shahkhali, Eric Yu, Farnaz Asayesh, Jamil Ahmad, Meron Teferra, Isabelle Arnulf, Pauline Dodet, Yo-El Ju, Michele T.M. Hu, Jacques Y. Montplaisir, Jean-François Gagnon, Alex Desautels, Abubaker Ibrahim, Ambra Stefani, Birgit Högl, Merve Akrtan-Süzgün, Alex Iranzo, Mónica Serradell, Angelica Montini, Gerard Maya, Carles Gaig, Gian Luigi Gigli, Mariarosaria Valente, Francesco Janes, Andrea Bernardini, Yves Dauvilliers, Karel Sonka, David Kemlink, Petr Dusek, Michael Sommerauer, Gültekin Tamgüney, Michela Figorilli, Monica Puligheddu, Valerie Cochen De Cock, Wolfgang Oertel, Annette Janzen, Elena Antelmi, Brit Mollenhauer, Claudia Trenkwalder, Friederike Sixel-Doring, Michele Terzaghi, Giuseppe Fiamingo, Matej Skorvanek, Kristina Kulcsarova, Beatriz Abril, Christelle Charley Monaca, Luigi Ferini-Strambi, Jitka Bušková, Beatrice Orso, Pietro Mattioli, Dario Arnaldi, Femke Dijkstra, Mineke Viaene, Bradley F. Boeve, Owen A. Ross, Guy A. Rouleau, Lee E. Neilson, Jonathan E. Elliott, Miranda M. Lim, Ronald B. Postuma, Ziv Gan-Or

**Author notes:** Corresponding author: Ziv Gan-Or, Department of Neurology and Neurosurgery McGill University, 1033 Pine Avenue West, Ludmer Pavilion, Room 312, Montréal, QC, Canada, H3A 1A1.

## Abstract

**Objective:** To explore potential genetic and/or causal associations between Post-Traumatic Stress Disorder and neurodegeneration-related isolated/idiopathic rapid-eye-movement sleep behavior disorder.

**Methods:** We conducted polygenic risk score, genetic correlation, and Mendelian randomization analyses using the latest genome-wide association studies summary statistics and individual genotyping data. Next, a blinded observer examined dopamine transporter imaging binding status—a marker of neurodegeneration—in patients with isolated/idiopathic rapid-eye movement sleep behavior disorder, with (N = 6) and without Post-Traumatic Stress Disorder (N = 32).

**Results:** Polygenic risk scores for Post-Traumatic Stress Disorder were associated with isolated/idiopathic rapid-eye-movement sleep behavior disorder, with each standard deviation increase linked to 14.7% higher odds (odds ratio = 1.15, 95% confidence interval: 1.04 to 1.26, p = 0.005). However, genetic correlation was weak, and Mendelian randomization did not support a potential causal relationship. The proportion of individuals with abnormal dopamine transporter imaging binding status was significantly higher in the Post-Traumatic Stress Disorder group compared to those without the disorder (p=0.01, X^2^ = 6.62).

**Interpretation:** Polygenic risk scores analysis identified an association between Post-Traumatic Stress Disorder and neurodegeneration-related isolated/idiopathic rapid-eye-movement sleep behavior disorder, consistent with the result from the small exploratory substudy. The lack of strong genetic correlation or causation may reflect limited sample size. Further research with larger and more diverse cohorts is crucial to clarify the genetic, biological and physiological mechanisms underlying this association.

## Introduction

Isolated/idiopathic rapid eye movement (REM) sleep behavior disorder (iRBD) is a parasomnia characterized by the lack of normal muscle paralysis during REM sleep, leading individuals to act out their dreams. Symptoms can vary from non-violent expressions like laughing and crying to more aggressive actions such as kicking or punching, which can potentially cause harm to both the individual and their sleep partner. iRBD is also an early clinical indicator of a-synucleinopathies and over 80% of individuals with iRBD will eventually develop either Parkinson’s disease (PD), dementia with Lewy bodies, and more rarely, multiple system atrophy. This association highlights the importance of investigating iRBD in the context of neurodegeneration^1^.

Previous clinical reports suggest a potential association between iRBD and post-traumatic stress disorder (PTSD). A report showed an early association between PTSD and RBD, where 56% (n=14/27) of individuals with RBD had a history of PTSD^2^. This was later reinforced through a cohort study of 394 United States Veterans, reporting an age-adjusted odds ratio of 3.4 associating the presence of PTSD with RBD, corresponding to an overall frequency rate of 15-20%^3^. Similarly, a study found that compared to individuals who suffered from trauma but did not develop PTSD, individuals with PTSD had ∼10-15 fold risk for RBD^4^. In parallel, several prospective longitudinal or retrospective pseudolongitudinal cohort studies have identified links between PTSD and neurodegenerative disorders. One study found that people with PTSD had nearly twice the risk of developing dementia compared to those without PTSD^5^, while others showed an increased risk of PD later in life^6,7^. A case-control study found that people with RBD and a history of both traumatic brain injury and PTSD showed worse synucleinopathy-related neurological measures than those with only RBD, highlighting the close relationship between these conditions and the need for further investigation of their shared mechanisms^8^.

In this study, we first aimed to examine whether there is genetic evidence for the association between PTSD and iRBD. We conducted polygenic risk score (PRS), genetic correlation and Mendelian randomization (MR) analyses to investigate potential genetic and causal relationships between PTSD and iRBD. Next, we aimed to explore dopamine transporter single-photon emission computed tomography (DaT-SPECT) binding status, a marker of neurodegeneration, in a subset of iRBD participants with and without PTSD. A blinded observer evaluated DaT-SPECT binding status as an objective marker of nigrostriatal degeneration in iRBD in consecutive participants from a single site enriched for PTSD, and comparisons between iRBD participants with and without PTSD were made, adjusting for known confounders

## Methods

### Population

We used the most recently published genome-wide association studies (GWAS) summary statistics for PTSD (PGC-PTSD Freeze 3) of European ancestry (N cases = 137,136; N controls = 1,085,746)^9^, and GWAS summary statistics for iRBD (N cases = 1,061; N controls = 8,386)^10^ for MR and genetic correlation analyses. Additionally, we analyzed individual genotyping data from a total of 1,292 iRBD cases (80% men, average age at onset: 60 +/− 13 years standard deviation (SD)) and 67,150 controls (53% men, average age: 57 +/− 9 years SD) from four major cohorts, which were used for polygenic risk score (PRS) analysis. These cohorts included the McGill cohort (N cases = 1,292; N controls = 1,353), NeuroGenetics Research Consortium (NGRC; dbGap phs000196.v3.p1; N controls = 1,968)^11^, National Institute of Neurological Disorders and Stroke (NINDS) Genome-Wide genotyping in Parkinson’s Disease (dbGap phs000089.v4.p2; N controls = 790)^12^, and UK Biobank (UKB; N controls = 63,039, Table 1)^13^. Moreover, we explored with DaT-SPECT a sub-analysis of participants with iRBD (N = 6 with PTSD; N = 32 without PTSD). Included participants were recruited and enrolled (between 11/2020 and 7/2024) at a single site, VA Portland Health Care System, with data contributed to the North American Prodromal Synucleinopathy (NAPS) Consortium as well as banked into a local repository (VA MIRB #4086).iRBD, referring to those who were diagnosed with RBD before developing overt neurodegeneration, was diagnosed according to the International Classification of Sleep Disorders (2nd or 3rd Edition), including video polysomnography.

**Table 1.** Summary of Cohorts and Analytical Methods Used in the Study.

| Study/Cohort | Number of Cases | Number of Controls | Data Type | Analysis |
| --- | --- | --- | --- | --- |
| Post-Traumatic Stress Disorder (PTSD) GWAS | 137,136 | 1,085,746 | GWAS summary statistics | MR, genetic correlation, PRS analysis |
| Idiopathic REM sleep behavior disorder (iRBD) GWAS | 1,061 | 8,386 | GWAS summary statistics | MR, genetic correlation |
| McGill cohort | 1,292 | 1,353 | Genotyping data | PRS analysis |
| NeuroGenetics Research Consortium (NGRC) | - | 1,968 | Genotyping data | PRS analysis |
| National Institute of Neurological Disorders and Stroke (NINDS) | - | 790 | Genotyping data | PRS analysis |
| UK Biobank (UKB) | - | 63,039 | Genotyping data | PRS analysis |

GWAS: genome-wide association study; MR: Mendelian randomization; PRS: polygenic risk score

### Genetic analysis

Genotyping of the McGill cohort was performed on the OmniExpress or Neurobooster arrays according to the manufacturer’s protocols (Illumina Inc). Pre-imputation quality control for all individual and variant-level data, except for UKB data, was completed as previously described (https://github.com/neurogenetics/GWAS-pipeline), and imputation was performed using the TOPMed Imputation Server with the TOPMed reference panel r3 and default settings^14^. Imputed genotyping data for UKB controls were obtained from unrelated individuals of European ancestry (field 22006), randomly selected after excluding those diagnosed with mental and behavioral disorders (codes F00-F99) and nervous system diseases (codes G00-G99), based on International Classification of Diseases version-10 (ICD-10, field 41270). All samples were confirmed to be of European descent via principal component analysis (PCA), and post-imputation quality control was conducted using a standard protocol (https://choishingwan.github.io/PRS-Tutorial/target/) to prepare the data for PRS analysis^15^.

We computed PTSD PRS for each iRBD case and each control using PRS continuous shrinkage (PRS-CS)^16^. Variants were selected via PRS-CS based on PTSD GWAS summary statistics, RBD genotyping data, and a European Linkage Disequilibrium reference panel recommended by PRS-CS (https://github.com/getian107/PRScs). Posterior single-nucleotide polymorphism (SNP) effect sizes were estimated using a continuous shrinkage approach within a Bayesian framework implemented by PRS-CS auto. Individual-level polygenic scores were computed using PLINK/2.0 (www.cog-genomics.org/plink/2.0)^17^ and Python 3.8.10 by averaging variant effect sizes within each chromosome and subsequently aggregating these values across chromosomes for each sample. We tested the association between PTSD polygenic scores and iRBD using a logistic regression model, adjusting for age, sex, cohort, and the first five principal components, using R version 4.1.2.

To estimate the genetic correlation between PTSD and iRBD, we employed linkage disequilibrium score regression (LDSC)^18^. Summary statistics for each dataset were preprocessed using the LDSC munge_sumstats.py script with an LD score reference panel derived from 1000 Genomes Phase 3 EUR ancestry (https://zenodo.org/records/10515792), ensuring the inclusion of high-quality SNPs. Subsequently, we utilized the ldsc.py script with the --rg flag to compute genetic correlations, applying the same LD score reference and the --no- intercept flag, as our data had no sample overlap.

To investigate whether there is a causal link between PTSD and iRBD, we performed MR using TwoSampleMR package in R^19^. Genetic variants associated with PTSD (the exposure) that reached genome-wide significance (p ≤ 5e-08) were filtered from the z-score based meta-analysis of the PTSD GWAS. Clumping was performed locally using PLINK/1.9 (www.cog-genomics.org/plink/1.9/)^17^ and an LD reference panel from 1000 Genomes Phase 3 EUR (https://zenodo.org/records/10515792) to select the SNP instruments. PTSD and iRBD data were then harmonized and MR was done using TwoSampleMR package to evaluate causality. The analysis was repeated with iRBD as the exposure and PTSD as the outcome to assess the causal impact of iRBD-associated genetics instruments on PTSD.

### Dat-SPECT analysis

Finally, thirty-eight consecutive iRBD participants with viable DaT-SPECT were included in retrospective analysis with the following criteria for grouping: PTSD (determined via the PTSD checklist for the Diagnostic and Statistical Manual of Mental Disorders, Fifth Edition (PCL-5), following standard clinical criteria (i.e., total score >32)) or non-PTSD (total score ≤32)^20^. DaT-SPECT abnormal status was determined via visual inspection by a blinded observer, a board-certified Neurologist with fellowship subspecialization in Movement Disorders (LEN). Chi-square analysis of the distribution of DaT-SPECT abnormal status was performed comparing PTSD and non-PTSD groups. Multiple logistic regression modeling was applied to control for potential confounders (e.g., age, sex, and history of brain injury).

## Results

### PTSD PRS is associated with increased risk of iRBD

We examined the potential genetic association between PTSD PRS and iRBD using logistic regression model. Individuals with a higher genetic risk for PTSD were more likely to develop iRBD, as for each one SD increase in the standardized PTSD PRS, the odds of having iRBD increased by 14.7% (odds ratio (OR) = 1.15, 95% confidence interval (CI): 1.04 to 1.26, p = 0.005). This suggests that there is a significant association between genetic risk for PTSD and the risk of iRBD.

However, using LDSC, we found a weak and small positive genetic correlation of only 0.03 (standard error (SE) = 0.02) that was not statistically significant (p = 0.11) suggesting that the shared genetic background between PTSD and iRBD may be overall limited (Table 2). We performed MR analysis to investigate a potential causal relationship between PTSD and iRBD. Using an inverse variance weighted approach with 43 PTSD genetic instruments, we obtained a beta estimate of 0.09 (SE = 0.42, p = 0.830) in the PTSD-to-iRBD direction (Table 3, Fig 1). Furthermore, in the iRBD-to-PTSD direction, only two significant SNPs remained, limiting statistical power and precluding robust conclusions.

**Fig 1.**
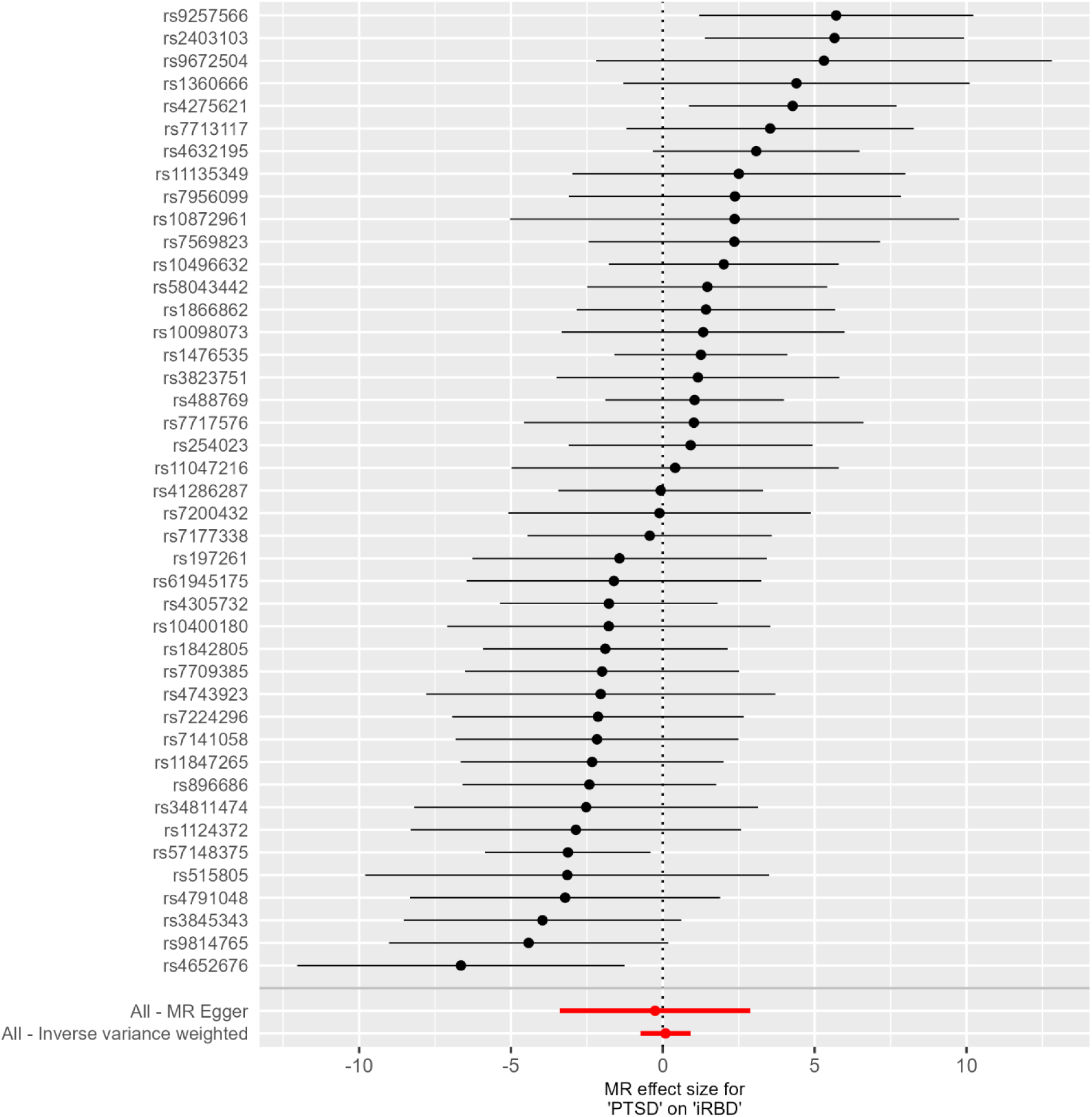
Mendelian Randomization Estimates for Post-Traumatic Stress Disorder (PTSD) on Isolated/idiopathic rapid-eye movement (REM) sleep behavior disorder (iRBD)

**Table 2.** Summary of Analytical Methods and Results.

| Analysis | Tool/Method | Estimate | Standard Error | P-value |
| --- | --- | --- | --- | --- |
| PRS Analysis | PRS-CS | $\beta$: 0.137 | 0.049 | 0.005 |
| Genetic Correlation | LDSC | rg: 0.031 | 0.019 | 0.106 |
| Mendelian Randomization | TwoSampleMR, IVW | $\beta$: 0.0907 | 0.422 | 0.830 |

PRS: polygenic risk score; PRS-CS: polygenic risk score with continuous shrinkage; LDSC: linkage disequilibrium score regression, a statistical method used to estimate genetic correlation from while accounting for linkage disequilibrium; IVW: inverse variance weighting, a Mendelian randomization method that combines effect estimates from multiple genetic variants; $\beta$ : beta coefficient, the estimated effect size in the statistical model; rg: genetic correlation, a measure of the shared genetic basis between two traits

**Table 3.** Mendelian Randomization Analysis.

| Method | N SNP | $\beta$ | SE | P-Value |
| --- | --- | --- | --- | --- |
| MR Egger | 43 | -0.2541169 | 1.5987146 | 0.8744881 |
| Weighted median | 43 | 0.340319 | 0.5242078 | 0.516205 |
| Inverse variance weighted | 43 | 0.0907476 | 0.422071 | 0.829763 |
| Simple mode | 43 | -2.0768048 | 1.3482727 | 0.1309776 |
| Weighted mode | 43 | 1.1385041 | 1.1450945 | 0.3257976 |

MR: Mendelian randomization; N SNP: number of single nucleotide polymorphisms used in the analysis; $\beta$ : beta coefficient, the estimated effect size in the statistical model; SE, standard error

### Individuals with iRBD and PTSD have higher rates of abnormal DaT-SPECT

Further exploring the association between PTSD, RBD, and potential increased risk of neurodegeneration, we performed a sub-analysis of neuroimaging data from iRBD participants co-enrolled in the NAPS Consortium, from a single site enriched for PTSD Of n=38 participants with RBD who underwent DaT-SPECT, there was a significant increase (p=0.01, X^2^ = 6.62) in the ratio of an abnormal DaT-SPECT status in those with PTSD (n=5/6 with PTSD showed abnormal DaT-SPECT; 83%) compared to those without PTSD (n=9/32 without PTSD showed abnormal DaT-SPECT; 28%). This significant association was maintained after correcting for age, sex, and history of brain injury via multiple logistic regression.

## Discussion

PRS analysis demonstrated a positive genetic association between PTSD PRS and the risk of developing iRBD. In other words, individuals with higher genetic risk for PTSD also have higher risk for iRBD. However, genetic correlation analysis did not indicate a strong shared genetic architecture, and no evidence of a potential causal relationship was observed using MR. The absence of a genetic correlation and causal link between PTSD and iRBD may be due to the limited power stemming from the small sample size of the iRBD GWAS. Identifying most genetic variants in highly polygenic traits requires large sample sizes to achieve genome-wide significance and capture the majority of genetic variance. Additionally, MR, which depends on a restricted set of significant variants to infer causality, faces challenges in identifying strong and valid instruments for polygenic traits. In contrast, PRS analysis is particularly powerful for detecting associations between polygenic traits by aggregating numerous SNP effects, even when individual effect sizes are small.

The relationship between PTSD, iRBD, and neurodegeneration is supported by several key studies. In one study of 394 veterans, researchers used in-lab video-polysomnography (PSG) and self-reported dream enactment to find that the overall prevalence of iRBD was 15% among those with PTSD^3^. In comparison, the prevalence of iRBD in the general population is estimated to be approximately 1%^1^. Also, Veterans with PTSD were approximately 2.8 times more likely to develop RBD, with a 147% increase in the prevalence ratio after age adjustment^3^. Another study that included RBD patients who underwent PSG and took sleep-related questionnaires revealed that, after controlling for age and apnea-hypopnea index, PTSD was more strongly associated with RBD compared to the trauma-exposed group without PTSD^4^. Furthermore, in a sample of 27 RBD patients, it was found that 15 individuals (56%) also met the criteria for co-occurring PTSD^2^. A retrospective study found that patients with PTSD had significantly higher REM sleep without atonia (RSWA) across multiple metrics compared to controls (all p < 0.025)^21^.

Beyond iRBD, PTSD is also a recognized independent risk factor for neurodegenerative diseases, particularly PD and Alzheimer’s disease (AD). A large cohort study of 181,093 midlife veterans showed that those with PTSD were 2.31 times more likely to develop dementia, including all dementia subtypes such as AD and Lewy body dementia^5^. A longitudinal study found individuals with PTSD had a higher likelihood of developing PD (2% vs. 0.5%) and a shorter interval between PTSD diagnosis and PD development^6^. Another study reported that PTSD was associated with a 1.5- to 1.9-fold increased risk of PD over successive five-year periods^7^. A study of 24 individuals with both traumatic brain injury and PTSD (neurotrauma) and RBD showed significantly greater impairments in cognitive, motor, and autonomic function compared to 96 individuals with only RBD, suggesting the combination may be linked to a more advanced neurodegenerative process or altered neuropathogenic trajectory^8^. Importantly, known PD risk genes such as GBA1 and LRRK2 do not appear to be individually associated with PTSD^9^. This possible association between PTSD, RBD, and increased risk of neurodegeneration, regardless of genetic predisposition is further supported by our sub-analysis of NAPS Consortium participants with DaT-SPECT. PTSD was significantly associated with an increased rate of abnormal DaT-SPECT interpretations, even after correcting for age, sex, and history of brain injury. While based on a small sample size and still exploratory, these data are consistent with clinical evidence for accelerated or altered neuropathology in iRBD with PTSD.

Our study has several limitations. One major limitation is the small sample size for the iRBD GWAS, which may be the reason for the limited power in detecting genetic correlations and conducting MR analyses. Another limitation is that our analysis focuses exclusively on individuals of European ancestry, due to the lack of PSG-confirmed iRBD in numerous other populations. This narrow scope may restrict the generalizability of our findings to other populations. Therefore, a key for future research is to replicate these analyses using a larger iRBD GWAS to increase the study’s power and to include diverse genetic backgrounds. Expanding the sample size and diversity would enhance both the statistical power and the validity of the findings. By addressing these limitations, future studies can provide deeper insights into the complex relationship between PTSD and RBD. In addition, we did not have access to the individual level genotypes in the PTSD GWAS, therefore we could not perform an analysis examining iRBD PRS effect on PTSD risk. Because DaT-SPECT is laborious and presents high burden and cost to both participants and studies, sample sizes will always be limited and additional work is needed that may also include other quantitative neurodegenerative outcome measures.

To conclude, our study suggests that individuals who are genetically at risk for PTSD, are also at risk for iRBD, providing further support for previous clinical observations. Early screening of individuals with PTSD for iRBD is therefore important, especially when clinical trials to prevent neurodegeneration and preventative treatment become available.

## Data Availability

The iRBD summary statistics are available on GWAS catalog (https://www.ebi.ac.uk/gwas/, study accession GCST90204200). PTSD summary statistics are publicly available on PGC website (https://pgc.unc.edu/for-researchers/download-results/ , accession ID ptsd2024). The UKB genotyping data were accessed through NeuroHub (https://www.mcgill.ca/hbhl/neurohub). Genotyping data from dbGaP (https://www.ncbi.nlm.nih.gov/gap/) are available through the NeuroGenetics Research Consortium (NGRC) (dbGaP accession: phs000196.v3.p1) and the National Institute of Neurological Disorders and Stroke (NINDS) Genome-Wide Genotyping in Parkinson's Disease study (dbGaP accession: phs000089.v4.p2). Due to the sensitive nature of potentially identifiable protected health information of participants, deidentified clinical data will be made available upon request pursuant to institutional approvals for a Data Use Agreement or equivalent agreement as appropriate. The code supporting the findings of this study will be made publicly available on GitHub upon publication at: https://github.com/mghamg/PTSD-RBD-Study

https://pgc.unc.edu/for-researchers/download-results/

https://www.mcgill.ca/hbhl/neurohub

https://www.ncbi.nlm.nih.gov/gap/

https://github.com/mghamg/PTSD-RBD-Study

## Acknowledgments

We would like to thank the research participants for contributing to this study. This study was financially supported through grants from the Hilary and Galen Weston Foundation, Michael J. Fox Foundation (MJFF) and the Canadian Consortium on Neurodegeneration in Aging (CCNA). Additionally, the G-Can (GBA1-Canada) Initiative, an open-science collaborative initiative aimed at addressing GBA1 mutation-based Parkinson’s disease, has made contributions to this research. G-Can is supported by The Hilary and Galen Weston Foundation, Silverstein Foundation, and J. Sebastian van Berkom and Ghislaine Saucier. This work was supported in part by the Intramural Research Program of the National Institute on Aging (NIA), and the Center for Alzheimer’s and Related Dementias (CARD), within the Intramural Research Program of the National Institute on Aging and the National Institute of Neurological Disorders and Stroke (1ZIAAG000546) as well as NIH NIA grants R34 AG056639, U19 AG071754, P50 AG016574, P30 AG062677 and VA RRD RX004822. M Skorvanek and KK received funding from the Slovak Research and Development Agency under contract no. APVV-22-0279, and the EU Renewal and Resilience Plan “Large projects for excellent researchers” under grant No. 09I03-03-V03-00007. The iRBD cohort study at First Medical Faculty (Prague, Czechia) is supported by the Czech Health Research Council (grant NU21-04-00535) and the National Institute for Neurological Research (Project No. LX22NPO5107), funded by the European Union – Next Generation EU. This research used the NeuroHub infrastructure and was undertaken thanks in part to funding from the Canada First Research Excellence Fund, awarded through the Healthy Brains, Healthy Lives initiative at McGill University, Calcul Québec and Compute Canada. This research has been conducted using the UK Biobank Resource under Application Number 45551. The UKB cohort was accessed using Neurohub (https://www.mcgill.ca/hbhl/neurohub). ZG-O is supported by the Fonds de recherche du Québec – Santé (FRQS) Chercheurs-boursiers award and is a William Dawson Scholar. MGS is supported by a graduate student award, Jeanne Timmins Costello Award. J.-F.G. holds a Canada Research Chair in Cognitive Decline in Pathological Aging. We acknowledge the contributions of Psychiatric Genomics Consortium Posttraumatic Stress Disorder Working Group (PGC-PTSD) for support with data sharing.

## Author Contributions

MGS conducted the analysis, interpreted the results, and wrote the manuscript. LL and MHGS assisted with computational analysis and support. EY performed quality control and imputation. FA, JA and M Teferra performed sample preparation and validation. IA, P Dodet, Y-EJ, MTMH, JYM, J-FG, AD, A Ibrahim, AS, BH, MA-S, A Iranzo, M Serradell, AM, GM, CG, GLG, M Valente, FJ, AB, YD, KS, DK, P Dusek, M Sommerauer, GT, MF, MP, VCC, WO, AJ, EA, BM, CT, FS-D, M Terzaghi, GF, M Skorvanek, KK, BA, CCM, LF-S, JB, BO, PM, DA, FD, M Viaene, BB, OR, GAR and RBP provided biospecimens and clinical data. LEN, JEE and MML contributed to biospecimens, clinical data, and neuroimaging data. ZG-O conceived the research idea, developed the methodology, supervised the study, and contributed to the interpretation of the results. All authors reviewed, approved and contributed to editing the final manuscript.

## Data Availability

The iRBD summary statistics are available on GWAS catalog (https://www.ebi.ac.uk/gwas/, study accession GCST90204200). PTSD summary statistics are publicly available on PGC website (https://pgc.unc.edu/for-researchers/download-results/ , accession ID ptsd2024). The UKB genotyping data were accessed through NeuroHub (https://www.mcgill.ca/hbhl/neurohub).

Genotyping data from dbGaP (https://www.ncbi.nlm.nih.gov/gap/) are available through the NeuroGenetics Research Consortium (NGRC) (dbGaP accession: phs000196.v3.p1) and the National Institute of Neurological Disorders and Stroke (NINDS) Genome-Wide Genotyping in Parkinson’s Disease study (dbGaP accession: phs000089.v4.p2). Due to the sensitive nature of potentially identifiable protected health information of participants, deidentified clinical data will be made available upon request pursuant to institutional approvals for a Data Use Agreement or equivalent agreement as appropriate. The code supporting the findings of this study will be made publicly available on GitHub upon publication at: https://github.com/mghamg/PTSD-RBD-Study

## References

1. Dauvilliers Y, Schenck CH, Postuma RB, et al. REM sleep behaviour disorder. Nat Rev Dis Primers 2018;4:19

2. Husain AM, Miller PP, Carwile ST. Rem sleep behavior disorder: potential relationship to post-traumatic stress disorder. J Clin Neurophysiol 2001;18:148–57

3. Elliott JE, Opel RA, Pleshakov D, et al. Posttraumatic stress disorder increases the odds of REM sleep behavior disorder and other parasomnias in Veterans with and without comorbid traumatic brain injury. Sleep 2020;43:

4. Lee E, Kim K, So HS, et al. REM Sleep Behavior Disorder among Veterans with and without Post-Traumatic Stress Disorder. Psychiatry Investig 2020;17:987–95

5. Yaffe K, Vittinghoff E, Lindquist K, et al. Posttraumatic stress disorder and risk of dementia among US veterans. Arch Gen Psychiatry 2010;67:608–13

6. Chan YE, Bai YM, Hsu JW, et al. Post-traumatic Stress Disorder and Risk of Parkinson Disease: A Nationwide Longitudinal Study. Am J Geriatr Psychiatry 2017;25:917–23

7. Scott GD, Neilson LE, Woltjer R, et al. Lifelong Association of Disorders Related to Military Trauma with Subsequent Parkinson’s Disease. Mov Disord 2023;38:1483–92

8. Elliott JE, Ligman BR, Bryant-Ekstrand MD, et al. Comorbid neurotrauma increases neurodegenerative-relevant cognitive, motor, and autonomic dysfunction in patients with rapid eye movement sleep behavior disorder: a substudy of the North American Prodromal Synucleinopathy Consortium. Sleep 2024;47:

9. Nievergelt CM, Maihofer AX, Atkinson EG, et al. Genome-wide association analyses identify 95 risk loci and provide insights into the neurobiology of post-traumatic stress disorder. Nat Genet 2024;56:792–808

10. Krohn L, Heilbron K, Blauwendraat C, et al. Genome-wide association study of REM sleep behavior disorder identifies polygenic risk and brain expression effects. Nat Commun 2022;13:7496

11. Hamza TH, Zabetian CP, Tenesa A, et al. Common genetic variation in the HLA region is associated with late-onset sporadic Parkinson’s disease. Nat Genet 2010;42:781–5

12. Simon-Sanchez J, Schulte C, Bras JM, et al. Genome-wide association study reveals genetic risk underlying Parkinson’s disease. Nat Genet 2009;41:1308–12

13. Bycroft C, Freeman C, Petkova D, et al. The UK Biobank resource with deep phenotyping and genomic data. Nature 2018;562:203–9

14. Taliun D, Harris DN, Kessler MD, et al. Sequencing of 53,831 diverse genomes from the NHLBI TOPMed Program. Nature 2021;590:290–9

15. Choi SW, Mak TS, O’Reilly PF. Tutorial: a guide to performing polygenic risk score analyses. Nat Protoc 2020;15:2759–72

16. Ge T, Chen CY, Ni Y, et al. Polygenic prediction via Bayesian regression and continuous shrinkage priors. Nat Commun 2019;10:1776

17. Chang CC, Chow CC, Tellier LC, et al. Second-generation PLINK: rising to the challenge of larger and richer datasets. Gigascience 2015;4:7

18. Bulik-Sullivan B, Finucane HK, Anttila V, et al. An atlas of genetic correlations across human diseases and traits. Nat Genet 2015;47:1236–41

19. Hartwig FP, Davies NM, Hemani G, et al. Two-sample Mendelian randomization: avoiding the downsides of a powerful, widely applicable but potentially fallible technique. Int J Epidemiol 2016;45:1717–26

20. Blevins CA, Weathers FW, Davis MT, et al. The Posttraumatic Stress Disorder Checklist for DSM-5 (PCL-5): Development and Initial Psychometric Evaluation. J Trauma Stress 2015;28:489–98

21. Feemster JC, Steele TA, Palermo KP, et al. Abnormal rapid eye movement sleep atonia control in chronic post-traumatic stress disorder. Sleep 2022;45:

